# A Double-Blind, Randomized, Controlled Phase III Trial Investigating Efficacy and Safety of Varenicline for Vaping Cessation in Adult Users

**DOI:** 10.1101/2022.12.20.22283715

**Authors:** Pasquale Caponnetto, Davide Campagna, Jasjit S. Ahluwalia, Chistopher Russell, Marilena Maglia, Paolo Marco Riela, Carmelo Fabio Longo, Barbara Busa’, Riccardo Polosa

**Affiliations:** Centre for the Prevention and Treatment of Tobacco Addiction (CPCT), University Teaching Hospital “Policlinico-Vittorio Emanuele”, University of Catania, Italy; Center of Excellence for the Acceleration of HArm Reduction (CoEHAR), University of Catania, Italy; UOC MCAU, University Teaching Hospital “Policlinico-Vittorio Emanuele”, University of Catania, Italy; Department of Clinical & Experimental Medicine, University of Catania, Italy; Brown University School of Public Health and Alpert School of Medicine, Providence, RI, USA; Russell Burnett Research & Consultancy Ltd, Glasgow, UK; Department of Mathematics and Informatics, University of Catania, Catania, Italy; UOC Farmacia Ospedaliera, ARNAS Hospital “Garibaldi”, Catania, Italy; ECLAT Srl, Spin-off of the University of Catania, Italy

**Author notes:** ***Corresponding author:*** Prof. Riccardo Polosa, Centro per la Prevenzione e Cura del Tabagismo (CPCT), Azienda Ospedaliero-Universitaria “Policlinico-V.Emanuele” dell’Università di Catania Via S. Sofia 78, 95123, Catania, Italy, *E-mail:.

**Keywords:** Vaping Cessation, e-cigarettes, varenicline, randomized controlled trial

## Abstract

**Background:** Vaping cessation is virtually unexplored. The efficacy and safety of varenicline for vaping cessation has not been studied and rigorous research is required to advance best practice and outcomes for e-cigarettes users who want to quit.

**Methods:** Eligible patients were randomized to either varenicline (1 mg, administered twice daily for 12 weeks) or placebo treatment (administered twice daily, for 12 weeks) combined with vaping cessation counseling. The trial consisted of a 12-week treatment phase followed by a 12-week follow-up, nontreatment phase. The primary efficacy endpoint of the study was biochemically validated continuous abstinence rate (CAR) at weeks 4 to 12. Secondary efficacy end points were the CAR at weeks 4 to 24 and 7-day point prevalence of vaping abstinence at weeks 12 and 24.

**Results:** CAR was significantly higher for varenicline vs placebo at each interval: weeks 4-12, 40.0% and 20.0%, respectively (OR = 2.67, 95% CI = [1.25 - 5.68], P = 0.011); weeks 4-24, 34.3% for varenicline and 17.2% for placebo (OR = 2.52, 95% CI = [1.14 - 5.58], P = 0.0224). The 7-day point prevalence of vaping abstinence was also higher for the varenicline than placebo at each time point. Serious adverse events were infrequent in both groups and not treatment-related.

**Conclusions:** Inclusion of varenicline and counseling in a vaping cessation program for EC users intending to quit may result in prolonged abstinence. These positive findings may also help guiding future recommendations for vaping cessation by health authorities and healthcare providers.

## INTRODUCTION

Electronic cigarettes (EC) are becoming increasingly popular with smokers worldwide (1-3). Users report buying them mainly to help abstain from smoking cigarettes, to relieve cigarette withdrawal symptoms, to save money, and to continue to have a ‘smoking’ experience but with reduced health risks (4,5).

Because ECs do not contain tobacco and do not rely on combustion to operate, the aerosol generated by ECs contain fewer and substantially lower level of harmful and potentially harmful chemicals compared to combustible tobacco cigarettes under normal conditions of use (6-8). For this reason, ECs have been proposed as a tool for reducing harm from cigarette smoking (9-11). Evidence from randomized controlled trials, observational studies, and population data converge on showing that EC use (‘vaping’) is an effective method of smoking cessation, with daily vaping being more effective than less frequent use (12-15). Nonetheless, the long-term health effects of combustion-free nicotine products are still not fully known and require investigation. The potential health impact of ECs has been addressed in two recent review articles, with conflicting conclusions (16,17). Moreover, perceptions of ECs being equally or more harmful than combustible cigarettes have increased in the past few years, raising concern among EC users about the potential health risks of vaping (18,19).

In addition to increasing concerns about the potential health risks of vaping, growing interest in quitting vaping has also been linked to the experience of some adverse physical effects (e.g., dry mouth, cough), the rising cost of vaping, and the need to break dependence on vaping products (20–22).

Although guidelines on best management for the cessation of combustible cigarettes are available (23,24), there are no evidence-based recommendations to assist ECs users intending to quit vaping, and it is unclear whether smoking cessation guidelines can be extrapolated to vaping products. In particular, there are no studies of the efficacy of medications approved for smoking cessation by the U.S. Food and Drug Administration (FDA) for aiding vaping cessation. Randomized controlled trials indicate that varenicline - a partial, high-affinity α4β2 nicotine receptor agonist - is more efficacious than placebo, bupropion (25,26), and nicotine replacement therapies (NRTs) for smoking cessation (27). The efficacy and safety of varenicline for vaping cessation has not been studied and rigorous research is required to guide the decisions of health authorities and healthcare providers.

The aim of this double-blind randomized placebo-controlled trial was to evaluate the efficacy and safety of varenicline (1 mg BID, administered for 12 weeks, and followed to week 24) combined with vaping cessation counseling in exclusive daily EC users intending to quit vaping. We also examined a number of variables that could have modulated the efficacy of the intervention in term of vaping abstinence.

## METHODS

### Participants

Exclusive EC users who were vaping daily and intending to quit vaping were screened for inclusion in this study.

Inclusion criteria were: (a) ≥ 18 years of age,; (b) exclusive daily EC use for ≥ 12 months; (c) at least one serious quit vaping attempt (defined as complete abstinence for at least 24 h) in the past; (d) willingness to quit vaping, confirmed by a ‘‘YES’’ response to each of two questions ‘‘Do you plan to quit vaping within the next 30 days?’’ and ‘‘Do you wish to participate in a vaping cessation program?’’; (e) self-reported reduction in vape consumption by at least 50% before committing to TQD (this instruction is given at screening).

Exclusion criteria were: (a) current diagnosis of mental illnesses including major depression, psychosis, or bipolar disorder that were diagnosed and treated by psychiatrists or clinical psychologists; (b) history of alcoholism or drug/chemical abuse within 12 months prior to screening; (c) known medical condition that, in the opinion of the investigators, would compromise subjects’ safety or participation; (d) currently pregnant or breast feeding or intending to become pregnant during the trial; e) use of vaping products containing zero nicotine.

Eligible subjects were recruited from local vape shops, databases of former smokers who attended a local smoking cessation center (Centro per la Prevenzione e Cura del Tabagismo - CPCT, University of Catania) and stopped smoking by switching to ECs, databases of former smokers who took part in CoEHAR (CoEHAR, University of Catania) sponsored tobacco harm reduction and switching studies, social networks, WhatsApp chat of the students of the University of Catania, and word of mouth among relatives and friends of study participants.

Eligible subjects were randomized to receive the active drug or placebo. The flow diagram of subjects is shown in **Figure 1**, according to the Consolidated Standards of Reporting Trials (CONSORT) reporting guideline. he ethical review board of the leading site, ienda spedaliero Universitaria Policlinico-Vi orio Emanuele, Università di Catania, reviewed and approved this RCT (approval reference number: n.88/2016/PO, 11/07/2016). All participants provided written informed consent. The study has been registered in EUDRACT with Trial registration ID: 2016-000339-42 (https://www.clinicaltrialsregister.eu/ctr-search/search?query=varevape).

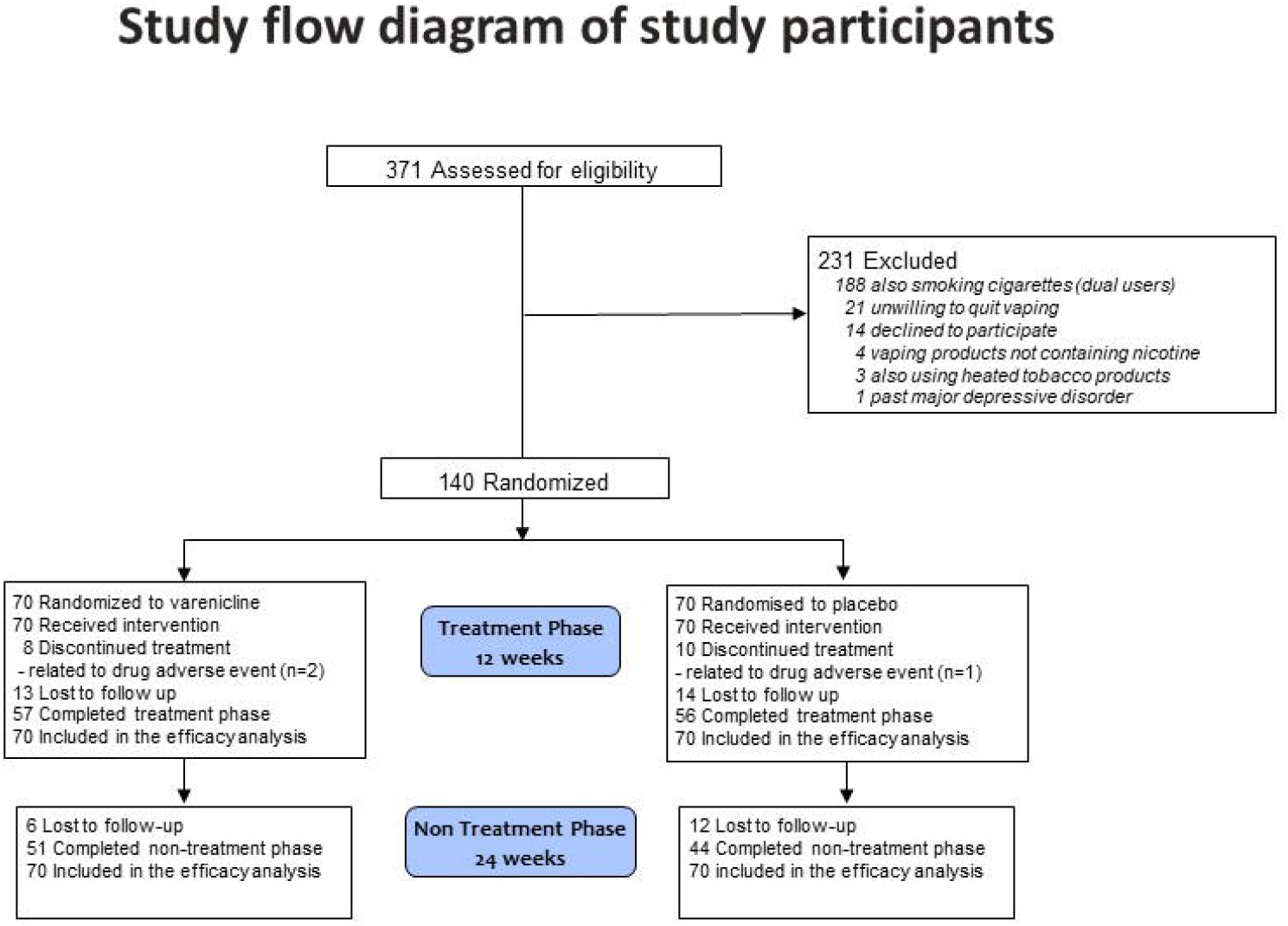

### Study Design

This was a double-blind, randomized controlled trial (RCT) of the efficacy and safety of combination varenicline (1 mg, administered twice daily for 12 weeks) plus counselling versus placebo (administered twice daily, for 12 weeks) plus counselling for vaping cessation in exclusive EC users who were vaping daily and intending to quit vaping. The total duration of the trial was 24 weeks, comprised of a 12-week treatment phase directly followed by a 12-week non-treatment phase (**Figure 2**). The study took place at Centro per la Prevenzione e Cura del Tabagismo (CPCT), the University-run smoking cessation center. The CPCT multidisciplinary team includes respiratory physicians, clinical psychologists, and professional nurses with more than 10 years’ experience in smoking cessation and e-cigarette management.

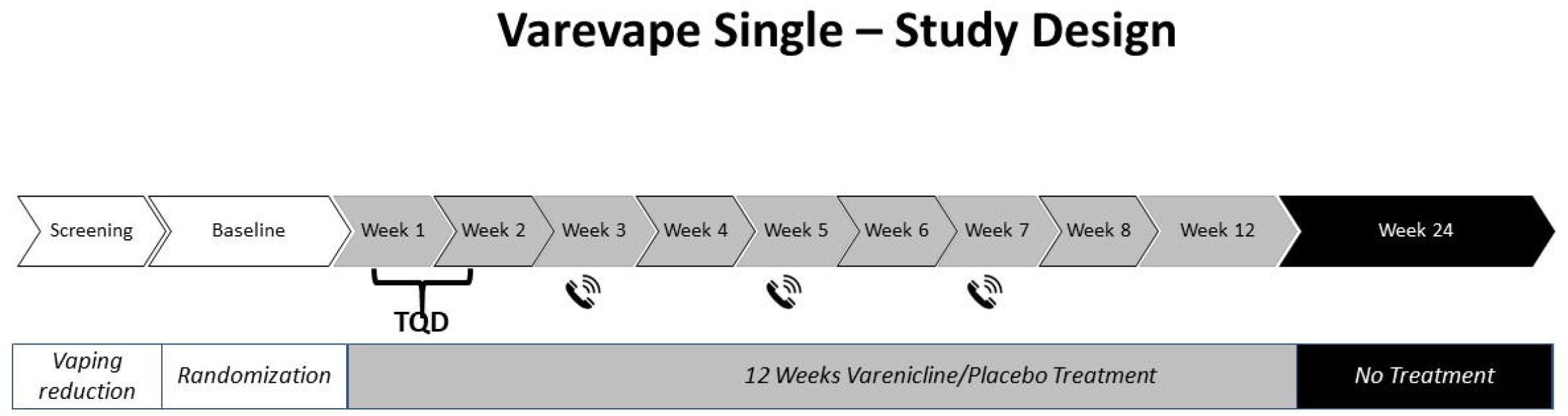

At screening, subjects who reported vaping daily and an intention to quit vaping were assessed for eligibility. Before leaving they were instructed to reduce the average daily vape use by at least 50% before making an appointment to attend the baseline visit (V1). At the baseline visit (V1), eligibility criteria were reassessed and subjects were randomized (1:1) to either varenicline plus vaping cessation counseling or placebo plus vaping cessation counseling. The list for treatment randomization was generated using SAS software (SAS Institute). The size of the blocks was a variable of 5, and the sequence of blocks was randomized and blinded. The following data were recorded at V1: sociodemographic characteristics, medical history, smoking and vaping history (including EC type, e-liquid flavor and nicotine concentration), and motivation (and reasons) for quitting vaping, vaping/nicotine consumption (assessed by modified Nicotine Use Inventory, mNUI), exhaled carbon monoxide (eCO) levels, blood pressure, heart rate, weight/Body Mass Index (BMI), questionnaires’ scores (Penn State Electronic Cigarette Dependence Index - PSECDI); Beck Depression Inventory-II - BDI-II); Beck Anxiety Inventory - BAI); Minnesota Nicotine Withdrawal Scale - MNWS), level of motivation to quit vaping (assessed by visual analogue score - VAS), and adverse events. Participants received their first vaping cessation counselling session and were instructed to set a target quit date (TQD) that was within the next 10 days. Prior to check-out, subjects were given a full week’s supply of the assigned treatment (either varenicline or placebo, depending on treatment arm). Study drugs were dispensed in accordance with the plan (**Table 1**). After V1, subjects were invited to return to the clinic on a weekly basis for the following 12 weeks (V2-V10), except for Visits 4, 6, and 8 (telephone contact). Subjects attending their TQD visit were required to have abstained or substantially reduced their vaping consumption. At each visit, subjects underwent vaping cessation counseling. Modified NUI, eCO levels, blood pressure, heart rate, weight/BMI (only at Week-12 visit), MNWS (only at Week-1, -2, - 4, -6, -8, -12 visit), and adverse events were recorded in the CRF at each study visit. At week-4 (V5), week-6 (V7), week-8 (V9) and week-12 (V10), saliva samples were collected for cotinine assessment. Study drugs were dispensed before check-out in accordance with the plan (**Table 1**). The study was continued in the non-treatment follow-up phase after completion of the treatment phase, consisting of a clinic visit at week-24 (V11). Modified NUI, eCO levels, blood pressure, heart rate, weight/BMI and MNWS were recorded in the CRF at this study visit. Collection of saliva samples was repeated.

**Table 1.**
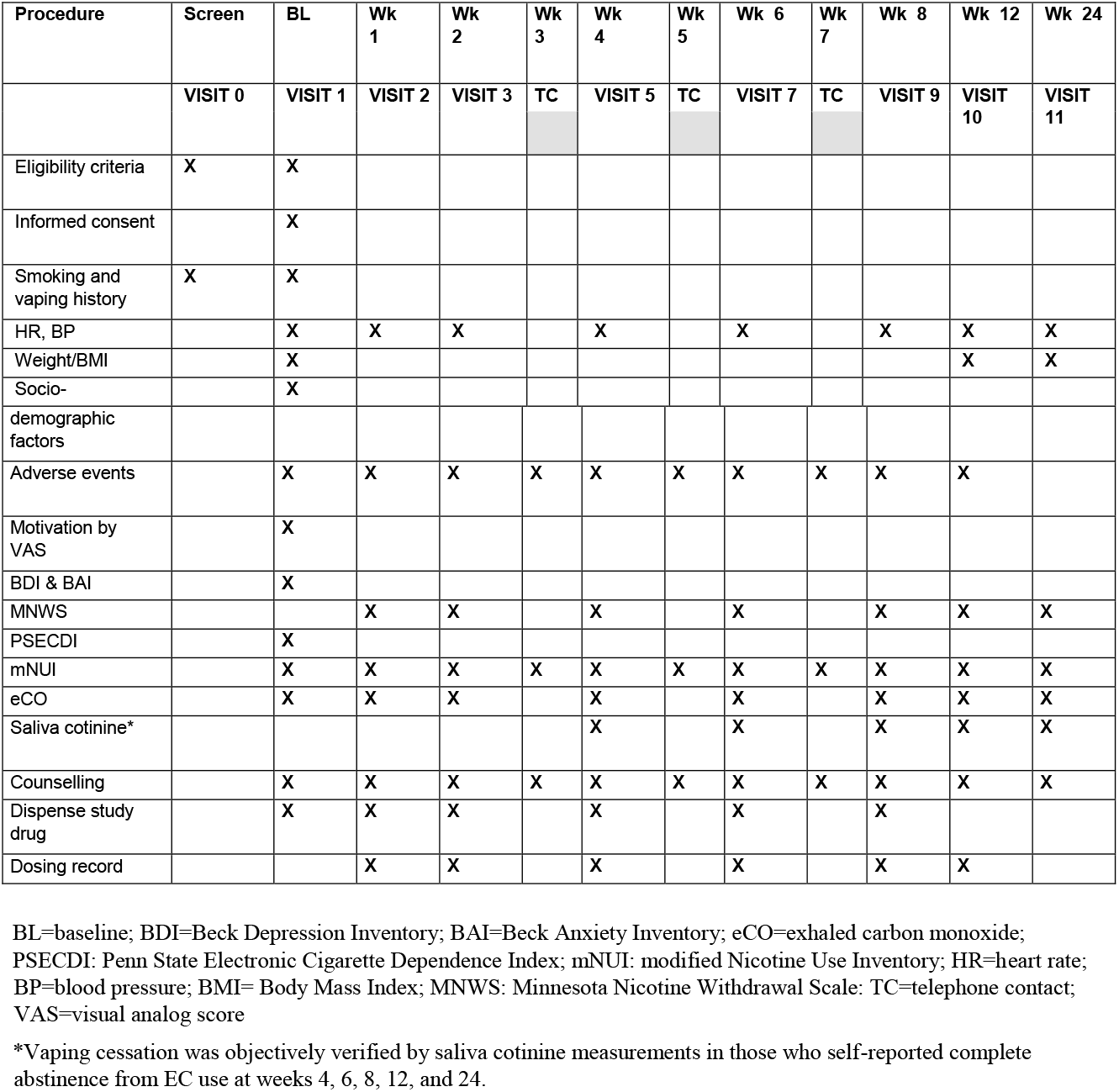
Study Schedule/Assessments.

| Procedure | Screen | BL | Wk 1 | Wk 2 | Wk 3 | Wk 4 | Wk 5 | Wk 6 | Wk 7 | Wk 8 | Wk 12 | Wk 24 |
| --- | --- | --- | --- | --- | --- | --- | --- | --- | --- | --- | --- | --- |
|  | VISIT 0 | VISIT 1 | VISIT 2 | VISIT 3 | TC | VISIT 5 | TC | VISIT 7 | TC | VISIT 9 | VISIT 10 | VISIT 11 |
| Eligibility criteria | X | X |  |  |  |  |  |  |  |  |  |  |
| Informed consent |  | X |  |  |  |  |  |  |  |  |  |  |
| Smoking and vaping history | X | X |  |  |  |  |  |  |  |  |  |  |
| HR, BP |  | X | X | X |  | X |  | X |  | X | X | X |
| Weight/BMI |  | X |  |  |  |  |  |  |  |  | X | X |
| Socio-demographic factors |  | X |  |  |  |  |  |  |  |  |  |  |
| Adverse events |  | X | X | X | X | X | X | X | X | X | X |  |
| Motivation by VAS |  | X |  |  |  |  |  |  |  |  |  |  |
| BDI & BAI |  | X |  |  |  |  |  |  |  |  |  |  |
| MNWS |  |  | X | X |  | X |  | X |  | X | X | X |
| PSECDI |  | X |  |  |  |  |  |  |  |  |  |  |
| mNUI |  | X | X | X | X | X | X | X | X | X | X | X |
| eCO |  | X | X | X |  | X |  | X |  | X | X | X |
| Saliva cotinine* |  |  |  |  |  | X |  | X |  | X | X | X |
| Counseling |  | X | X | X | X | X | X | X | X | X | X | X |
| Dispense study drug |  | X | X | X |  | X |  | X |  | X |  |  |
| Dosing record |  |  | X | X |  | X |  | X |  | X | X |  |

BL=baseline; BDI=Beck Depression Inventory; BAI=Beck Anxiety Inventory; eCO=exhaled carbon monoxide; PSECDI: Penn State Electronic Cigarette Dependence Index; mNUI: modified Nicotine Use Inventory; HR=heart rate; BP=blood pressure; BMI= Body Mass Index; MNWS: Minnesota Nicotine Withdrawal Scale; TC=telephone contact; VAS=visual analog score
\*Vaping cessation was objectively verified by saliva cotinine measurements in those who self-reported complete abstinence from EC use at weeks 4, 6, 8, 12, and 24.

### Study Outcomes Measures

The primary efficacy endpoint of the study was the proportion of subjects with continuous abstinence from vaping between week 4 to week 12 (CAR 4-12 Wks). Abstinence from vaping was defined as cotinine-verified (saliva cotinine < 10 ng/ml) self-reported abstinence from EC use since last study visit. This definition of abstinence also includes eCO-verified (□ 7 ppm) self-reported abstinence from tobacco smoking (in case of relapse back into cigarette smoking). Secondary efficacy endpoints were CAR 4-24 Wks, and 7-day point prevalence of abstinence at weeks 12, and 24.

Safety endpoints included information on the number of adverse events (AE), and serious adverse events (SAE) occurring between treatment randomization (V1) and last week of treatment (V10). Between and within treatment groups changes were reported for blood pressure, heart rate, weight, and BMI.

Secondary analyses by vaping phenotype classification (continuous quitters vs. treatment failures, which includes anything that does not fall under CAR; i.e., reducers, relapsers, and lost to follow up) will be reported in separate papers.

### Trial Interventions

#### Vape reduction

A reduction of at least 50% in daily vaping was targeted as a preparatory strategy. Instructions about vaping reduction were specific to the type of vaping product used. If it was a refillable device, the 50% reduction was indicated by a reduction in e-liquid volume consumed per day (e.g. reducing down to at least 2 ml/day if he/she vaped on average 4 ml/day). If it was a closed system (e.g. prefilled pod/cartridge) the 50% reduction was indicated by doubling the number of days over which the same volume was consumed (e.g. reducing down to at least 1 pod every two days if he/she consumed 1pod/day). Potential study subjects were instructed to gradually taper down daily consumption at their own pace, over time.

#### Cessation Medications

Varenicline (0.5 mg tablet) and matched placebo tablets were supplied by the study sponsor randomized and distributed by the hospital pharmacy. Blinding was ensured by the identical appearance of drug and placebo tables and their containers. Subjects assigned to varenicline were titrated to full dose by the time of their TQD (0.5 mg/day for 2-3 days, 0.5 mg twice daily for 4-5 days; then 1 mg twice daily for 11 weeks), according to manufacturer’s recommendations.

#### Vaping cessation counselling

Subjects in both treatment groups received the same vaping cessation counselling throughout the whole duration of the study. One-on-one counselling were provided at each visit for a total of 1015 minutes by two experienced clinical psychologists.This behavioral intervention is described in detail in the Appendix 1. Briefly, our approach to vaping cessation was partially adapted from the 5 ‘s brief tobacco interventions for smokers who are ready to quit (28). <u>First</u>, we collected information about participants’ frequency and intensity of use of vaping products (at baseline).

<u>Second</u>, we **assessed** readiness to quit vaping by asking two questions: ‘‘Do you plan to quit vaping within the next 30 days?’’, ‘‘Do you wish to participate in a vaping cessation program?’’ Before attending the baseline visit (and committing to a target quit date - TQD), potential study subjects were asked to reduce the daily use/consumption of their vaping product by at least 50%. When vaping frequency was reduced by 50% (indicating readiness to commit to vaping cessation plan and TQD), they were admitted to the baseline visit. <u>Third</u>, those who successfully reduced by 50% their daily use were **assisted** with a quit plan (combining vaping reduction, cessation counseling, use of varenicline, and close follow ups). Participants were instructed to set a TQD, ideally within two weeks. Participants were reminded of the challenges posed by craving and nicotine withdrawal symptoms when stopping vaping products completely and counseled on how to cope with them to avoid a relapse to vaping (or worse to smoking). Close follow up in the first four weeks of the cessation program was arranged to assess participants’ progress, review stress coping skills in order to mitigate the possibility of vaping relapse, address varenicline’s adverse events, and maintain participants’ motivation to quit. Participants were assisted ***in dealing with***

#### cravings and withdrawal

As nicotine is an important determinant of e-cigarette dependence, the withdrawal effects experienced upon cessation of vaping may be similar in nature, frequency, and intensity to those experienced when trying to quit tobacco cigarettes. Therefore, vaping cessation counselling may usefully adapt strategies that have long been trained as part of behavioral counselling for smoking cessation. Practical counseling in this study focused on two elements:

1. Helping the participant to identify situations that have historically triggered the individual’s motivation to vape (e.g., social situations, stressful situations, negative emotions).
2. Assisting the participant to practice using a range of cognitive and behavioral coping skills in response to trigger situations.

### Study Assessments

The activities carried out during study visits are listed (**Table 1**) and included the following measurements: 1) resting heart rate (HR), systolic (SBP) and diastolic (DBP) blood pressure taken with a semi-automated oscillometric sphygmomanometer (Smart Pressure, CA-MI Snc, Parma, Italy); 2) body weight and height taken with a mechanical column scale and standing scale slide bar (Seca, Intermed Srl, San Giuliano Milanese, Italy); 3) Body Mass Index (BMI) calculated by dividing weight by height^2^ (kg/m^2^); 4) exhaled carbon monoxide (eCO) levels evaluated with a calibrated handheld device (MicroCO, Vyare Medical Inc.); 5) Penn State Electronic Cigarette Dependence Index (PSECDI), a 10-item questionnaire used to measure e-cigarette dependency (29); 6) Beck Depression Inventory-II (BDI-II), a 21-item questionnaire used to measures subjective rating of depression; 7) Beck Anxiety Inventory (BAI), a 21-item questionnaire used to rate subjective physiological and cognitive symptoms associated with anxiety; 8) Minnesota Nicotine Withdrawal Scale (MNWS), a 9-item questionnaire used to evaluate nicotine withdrawal symptoms after cessation.

Saliva samples were collected for cotinine measurement in those who stated they had not vaped and with an eCO □ 7 ppm (just to confirm no combustible cigarette use). Participants were asked to chew a small cotton roll (TR0N00RU2, Dentalica, Milano, Italy) for 60 seconds. Cotton rolls were placed into polypropylene tubes and stored at - □ 20 C until use. Cotinine concentrations in saliva samples were analyzed in duplicate by gas chromatography (30). We adopted a salivary cotinine cut-off for abstinence of 10 ng/ml (31,32).

### Safety reporting

Safety data were summarized for both treatment groups and summary statistics reported. Any events documented in the period from the point of treatment initiation until last week of treatment (week-12, V10) was considered relevant to the safety analysis.

Adverse events: all observed or volunteered AEs, regardless of treatment group or suspected causal relationship to study drug, were recorded. Events involving adverse drug reactions, illnesses with onset during the study were recorded. For all AEs, sufficient information was obtained by the investigator to determine the causality of the AEs.

Serious adverse events: all SAEs (as defined below) regardless of treatment group or suspected relationship to study drug were reported immediately. A SAE is any adverse drug experience occurring at any dose that: 1. Results in death; 2. Is life-threatening; 3. Results in inpatient hospitalization or prolongation of existing hospitalization; 4. Results in a persistent or significant disability/incapacity.

### Statistical Methods

No success rates on varenicline use among vapers was available to determine the correct sample size for this study, the first of its type. However, an RCT that involved 139 long-term NRT users to assess the impact of varenicline combined with counseling to help people quit NRT revealed a significant difference between the active vs. placebo arm (33). Consequently, a similar sample size of 140 participants was selected for this study.

Baseline and demographic data are listed for all treatment groups. Summary statistics are reported for each treatment group. At baseline, differences between the varenicline and placebo groups were evaluated by means of one-way analysis of variance (ANOVA) and Mann-Whitney U-test for normally and non-normally distributed continuous data, respectively; χ^2^ test was used to test differences on categorical variables. Secondary endpoints were analyzed using procedures similar to that described above for the primary endpoint. Intention-to-treat analysis were adopted for efficacy evaluation, on the assumption that subjects lost to follow-up continued vaping.

Safety data were summarized for both treatment groups and summary statistics reported. Any events documented in the period from the point of treatment initiation until last week of treatment (week-12, V10) was considered as relevant to the safety analysis.

Aimed at identifying possible predictors of continuous vaping abstinence, a multiple logistic regression model was estimated in which continuous abstinence between weeks 9-12 (yes/no) was entered as the criterion variable. Putative predictors, selected by *a priori* evaluation of the variables that could act as determinants of successful continuous vaping abstinence, were entered as independent variables which included baseline characteristics (age, gender, years of smoking prior to regular vaping, years of exclusive daily vaping, number of quit vaping attempts, motivation levels by VAS, cohabitant vapers, BDI II score, BAI score, PSECDI score, MNWS at week-4, weight increase at week-12, and study group) as covariates, estimated the varenicline group vs the placebo group that could act as determinants of successful continuous vaping abstinence, were entered as independent variables.

The analyses were performed using Python 3.6 with Pandas 1.3.5, SciPy 1.7.3 and Statsmodel 0.12.2, and jamovi 2.3.16. All tests were 2-sided, and P < 0.05 was considered to be significant.

## RESULTS

### Trial Participants

Of a total of 371 EC users screened for eligibility, 140 were randomized to either varenicline plus vaping cessation counseling (N = 70) or placebo plus vaping cessation counseling (N = 70). All EC users included in the study were former smokers. he flow diagram of study subjects’ participation in the trial is shown in **Figure 1**. Most screen failures were due to dual use and no desire to quit vaping. One hundred and thirteen participants completed all the visits within the treatment phase, of whom 57 were in the varenicline group and 56 in the placebo group. The 24week study visit (non-treatment phase) was completed by 95 subjects, of whom 51 were in the varenicline group and 44 were in the placebo group. Subjects’ baseline characteristics between groups were comparable with the exception of age, BAI, PSECDI, and educational level (**Table 2**). Subjects had a mean (SD) age of 52.6 (9.1) years, and smoked 15-20 cigarettes daily for at least 25 years before switching to vaping products. Participants were exclusive daily EC users for at least 2 years, had made at least one serious quit vaping attempt in the past, and had a mean (SD) PSECDI score of 11.7 (6.2) for the varenicline group and 14.9 (7.3) for the placebo group, indicating high level of EC dependency. Participants self-reported reduction in daily EC use by at least 50%, and had a VAS motivation score > 8, indicating strong motivation to quit vaping. Approximately 90% of study subjects used a refillable system as their main EC device (mainly, refillable tank systems) with tobacco and fruit flavored e-liquids as the most common vape products. The reasons most commonly endorsed for an interest in quitting vaping were: a) concern about the potential health risks of long-term EC use (72.9%) ; b) desire to break dependence on vaping products (62.1%); c) the experience of some adverse physical effects (e.g., dry mouth, sore throat, dry cough) (27.9%); and d) the increasing cost of vaping (23.6%).

**Table 2.**
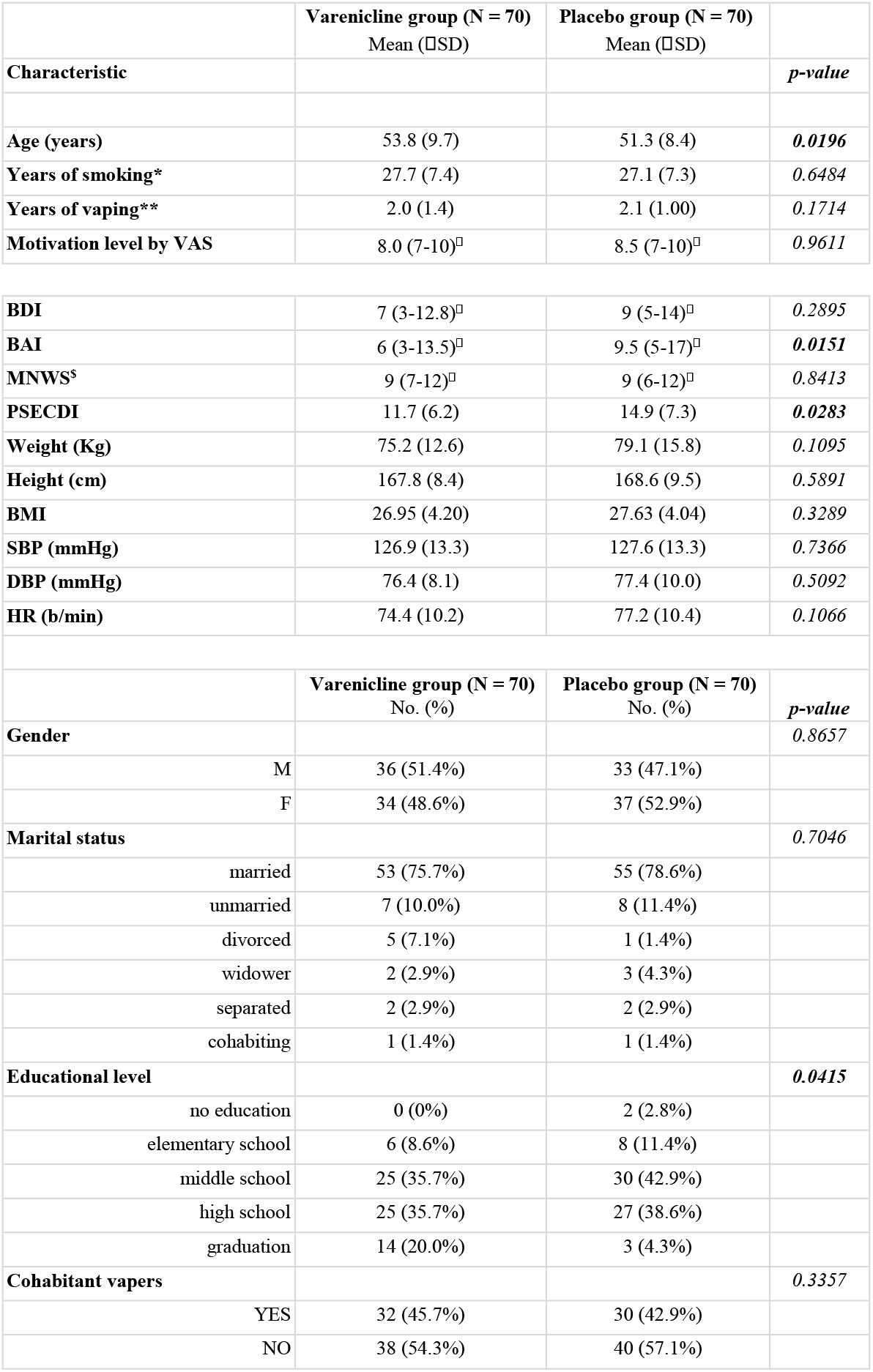
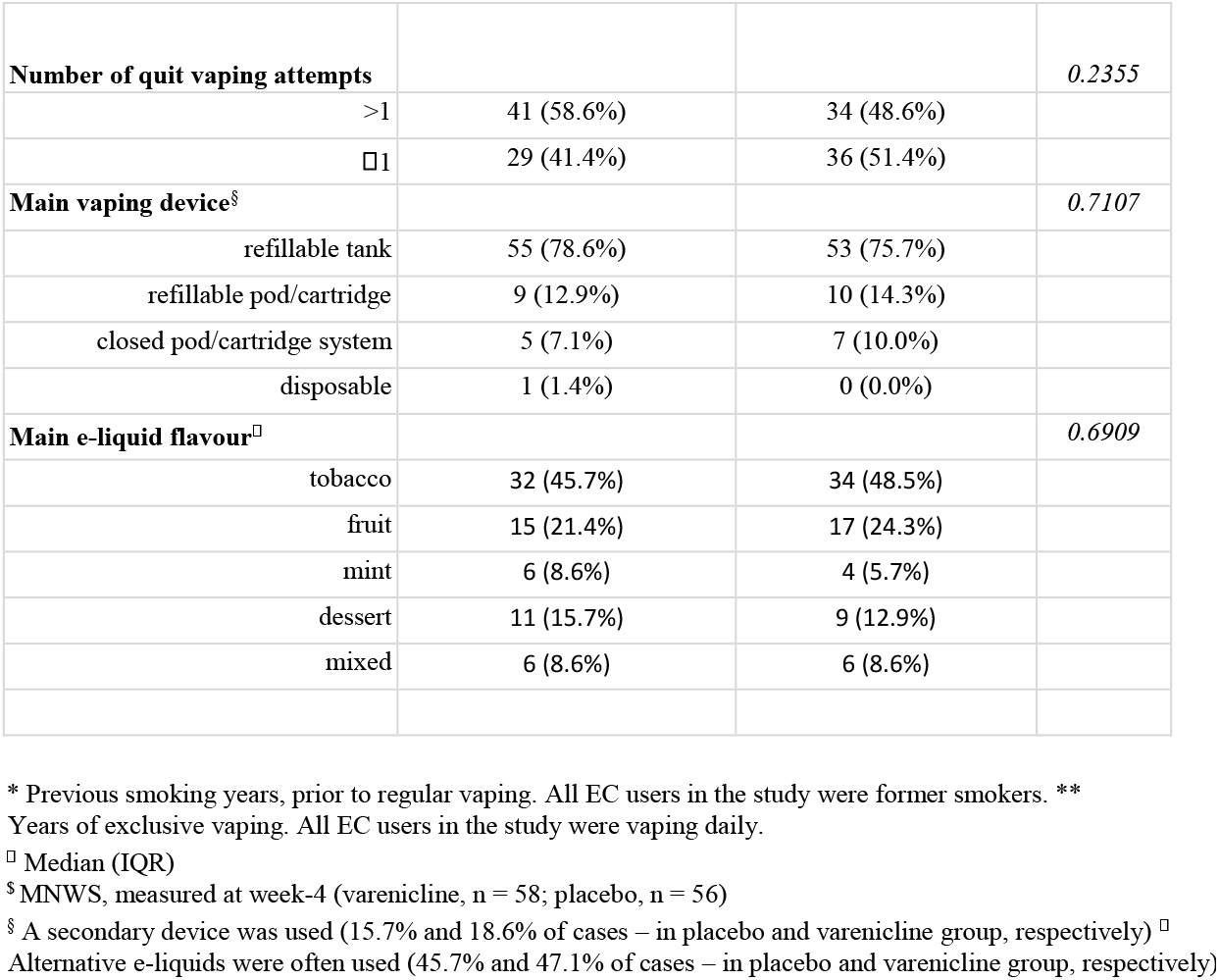
Baseline characteristics of study participants by treatment group.

### Vaping abstinence rates

The cotinine level-verified CARs for weeks 4-12 and weeks 4-24 are shown in **Figure 3 and Table 3**. CARs were significantly higher for varenicline vs placebo at each interval: weeks 4-12, 40.0% and 20.0%, respectively (OR = 2.67, 95% CI = [1.25 - 5.68], P = 0.011); weeks 4-24, 34.3% for varenicline and 17.2% for placebo (OR = 2.52, 95% CI = [1.14 - 5.58], P = 0.0224).

**Table 3.** – Change in vaping behavior* throughout the study.

|  | **Placebo Arm**<br>**N = 70** |  |  |  |  | **Varenicline Arm**<br>**N = 70** |  |  |  |  |
| --- | --- | --- | --- | --- | --- | --- | --- | --- | --- | --- |
|  | Total<br>Subjects<br>N | LTFU<br>N | Vape<br>Reducers<br>N | Vape<br>Relapsers<br>N | Vape<br>Quitters<br>N (%) | Total<br>Subjects<br>N | LTFU<br>N | Vape<br>Reducers<br>N | Vape<br>Relapsers<br>N | Vape<br>Quitters<br>N (%) |
| At week4 | 58 | 12 | 17 | 25 | 16<br>(22.9%) | 58 | 12 | 19 | 10 | 29 (41.4%) |
| At week5 | 58 | 12 | 15 | 24 | 19<br>(27.1%) | 57 | 13 | 17 | 11 | 29 (41.4%) |
| At week6 | 58 | 12 | 15 | 25 | 18<br>(25.7%) | 57 | 13 | 19 | 10 | 28 (40.0%) |
| At weeks-7 | 58 | 12 | 16 | 24 | 18<br>(25.7%) | 57 | 13 | 21 | 8 | 28 (40.0%) |
| At weeks-8 | 56 | 14 | 17 | 25 | 14<br>(20.0%) | 57 | 13 | 22 | 7 | 28 (40.0%) |
| At<br>week12 | 56 | 14 | 15 | 27 | 14<br>(20.0%) | 57 | 13 | 23 | 6 | 28 (40.0%) |
| At<br>week24 | 44 | 26 | 10 | 22 | 12<br>(17.1%) | 51 | 19 | 15 | 12 | 24 (34.3%) |

\*self-reported (cotinine-verified at 4-, 6-, 8-, 12-, and 24-weeks) LTFU – lost to follow up

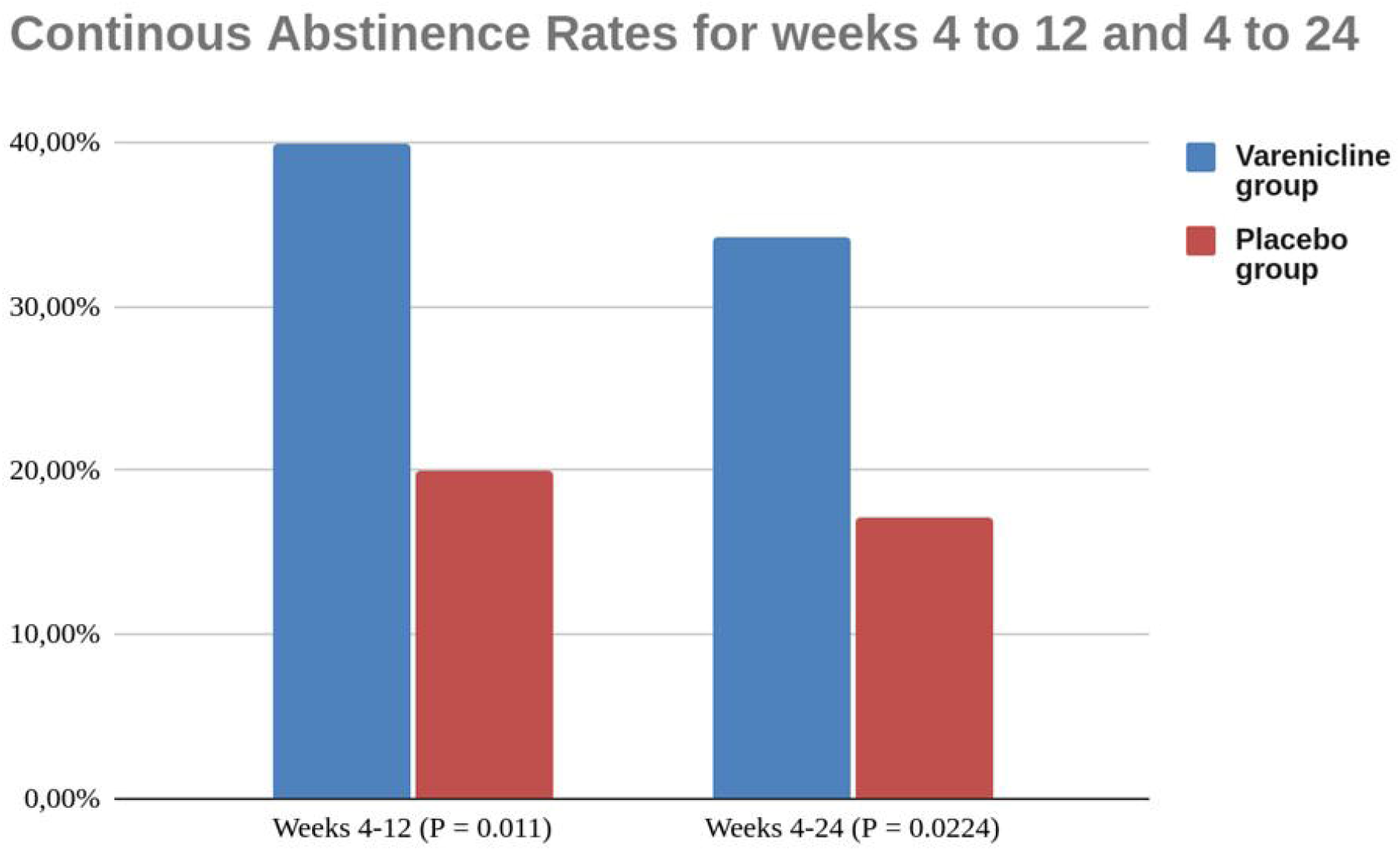

The 7-day point prevalence of vaping abstinence was also higher for the varenicline than placebo at each time point **Table 3**; in particular, significant results were shown at week 4, 41.4% vs 22.9% (OR = 2.39, 95% CI = [1.15 - 4.97], P = 0.02); week 12, 40.0% vs 20.0% (OR = 2.67, 95% CI = [1.25 - 5.68], P = 0.011); and week 24, 34.3 vs 17.2 (OR = 2.52, 95% CI = [1.14 - 5.58], P = 0.0224).

### Changes in vaping behavior

EC users who were not abstinent from vaping were considered as treatment failures. Treatment failures consisted of subjects who relapsed back into vaping, who were unable to completely stop vaping (i.e. reducers), and who were lost to follow-up. Taking the whole cohort of subjects completing the study, reduction in vaping consumption was observed in 33.6% and 26.3% of the subjects at week 12 and week 24, respectively. Vaping relapse was observed in 29.2% and 35.8% of the subjects at week 12 and week 24, respectively. No subject in the study relapsed to tobacco cigarette smoking. Details of changes in vaping status at each study visits are illustrated in **Table 4**.

**Table 4.** – Change in vaping behavior* throughout the study.

|  | Placebo Arm<br>N = 70 |  |  |  |  | Varenicline Arm<br>N = 70 |  |  |  |  |
| --- | --- | --- | --- | --- | --- | --- | --- | --- | --- | --- |
|  | Total<br>Subjects<br>N | LTFU<br>N | Vape<br>Reducers<br>N | Vape<br>Relapsers<br>N | Vape<br>Quitters<br>N (%) | Total<br>Subjects<br>N | LTFU<br>N | Vape<br>Reducers<br>N | Vape<br>Relapsers<br>N | Vape<br>Quitters<br>N (%) |
| At week4 | 58 | 12 | 17 | 25 | 16<br>(22.9%) | 58 | 12 | 19 | 10 | 29 (41.4%) |
| At week5 | 58 | 12 | 15 | 24 | 19<br>(27.1%) | 57 | 13 | 17 | 11 | 29 (41.4%) |
| At week6 | 58 | 12 | 15 | 25 | 18<br>(25.7%) | 57 | 13 | 19 | 10 | 28 (40.0%) |
| At weeks-7 | 58 | 12 | 16 | 24 | 18<br>(25.7%) | 57 | 13 | 21 | 8 | 28 (40.0%) |
| At weeks-8 | 56 | 14 | 17 | 25 | 14<br>(20.0%) | 57 | 13 | 22 | 7 | 28 (40.0%) |
| At<br>week12 | 56 | 14 | 15 | 27 | 14<br>(20.0%) | 57 | 13 | 23 | 6 | 28 (40.0%) |
| At<br>week24 | 44 | 26 | 10 | 22 | 12<br>(17.1%) | 51 | 19 | 15 | 12 | 24 (34.3%) |

\*self-reported (cotinine-verified at 4-, 6-, 8-, 12-, and 24-weeks) LTFU – lost to follow up

### Clinical and Demographic Features Influencing Vaping Abstinence

A multiple logistic regression model, which included baseline characteristics (age, gender, years of smoking prior to regular vaping, years of exclusive daily vaping, number of quit vaping attempts, motivation levels by VAS, cohabitant vapers, BDI II score, BAI score, PSECDI score, MNWS at week4, weight increase at week-12, and study group) as covariates, estimated the varenicline group vs the placebo group had an OR of 3.2 (95% CI, 1.19-8.60; *P* = .021) for the CAR at weeks 4 to 12 (**Table 5**). Having cohabitant vapers reduced the odds of success for CAR by approx. 70% (OR, 0.284; 95% CI, 0.091-0.888; *P* = .030). High BAI scores were also associated with reduced odds of success for CAR (OR, 0.219; 95% CI, 0.055-0.870; *P* = .031). Non-significant trends were observed for PSECDI (p = .054) and craving score (p = .059).

**TABLE 5.**
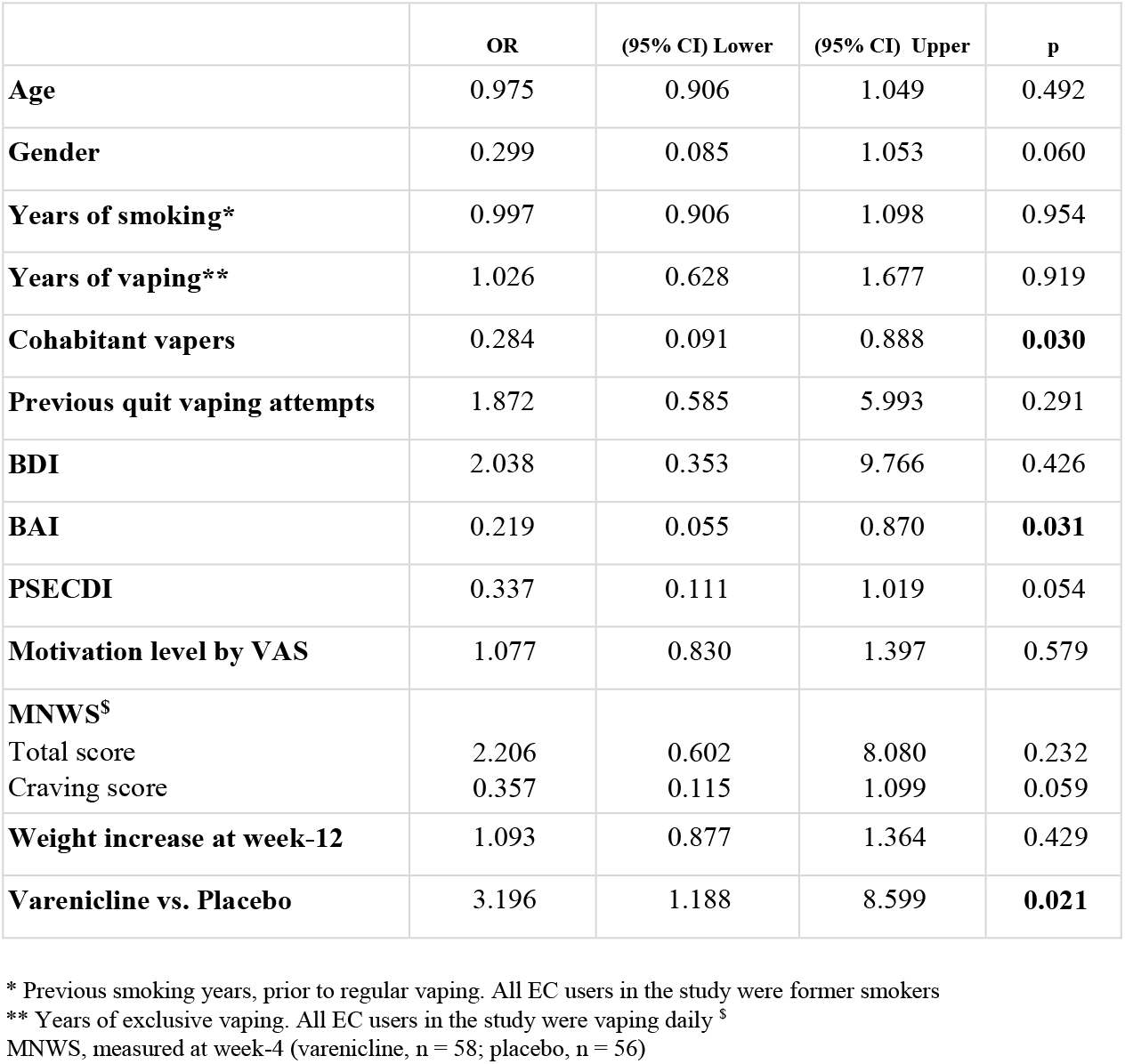
- Multivariate Logistic Regression Model for CAR 4-12 Week.

### Adverse Events

A total of 36 subjects (51.4%) who received varenicline and 29 patients (41.4%) who received placebo reported AEs. Most AEs were rated as mild or moderate and rarely led to treatment discontinuation; two in the varenicline group and one in the placebo group. The AEs that occurred more frequently in the varenicline group than in the placebo group were nausea (49 [19.9%] vs 19 [12.3%]), flatulence (17 [6.9%] vs 6 [3.9%]), and abnormal dreams (16 [6.5%] vs 5 [3.3%]) (**Table 6**).

**Table 6.** Summary of adverse events occurring in 3% or more of either treatment group.

|  | Varenicline<br>n(%) | Placebo<br>n(%) |
| --- | --- | --- |
| Adverse Events* |  |  |
| Total reported events | (N = 246) | (N = 154) |
| *Nausea* | 49 (19.9%) | 19 (12.3%) |
| *Headache/Migraine* | 25 (10.2%) | 16 (10.4%) |
| *Dry mouth* | 24 ( 9.8%) | 22 (14.3%) |
| *Insomnia* | 27 (11.0%) | 12 ( 7.8%) |
| *Flatulence* | 17 ( 6.9%) | 6 ( 3.9%) |
| *Abnormal dreams* | 16 ( 6.5%) | 5 ( 3.3%) |
| *Anxiety* | 16 ( 6.5%) | 9 ( 5.8%) |
| *Irritability* | 14 ( 5.7%) | 10 ( 6.5%) |
| *Sore throat* | 16 ( 6.5%) | 16 (10.4%) |
| *Constipation* | 10 ( 4.1%) | 9 ( 5.8%) |
| *Cough* | 10 ( 4.1%) | 9 ( 5.8%) |
| *Lightheadedness/dizziness* | 10 ( 4.1%) | 8 ( 5.2%) |
| *Others* | 12 ( 4.9%) | 13 ( 8.4%) |

\*Adverse events recorded during treatment phase and occurring in 3% or more of either treatment group. Participants were counted only once per row but could be counted several times per column.

Patients did not report suicidal ideation, worsening of depressive symptoms, or serious AEs (with the exception of one complication of inguinal hernial in the varenicline arm at week 12). No significant changes in mean (SD) systolic and diastolic blood pressure, and resting heart rate were observed between and within treatment groups (**Table 7 A,B**). High blood pressure was detected in 8 subjects (3, varenicline; 5 placebo). Weight gain was observed among subjects in the varenicline group; however, the increase of 1.4 kg and 2.2 kg at week-12 and week-24 was nonsignificant. The frequency of most commonly reported respiratory/oral AEs (such as dry mouth, sore throat, cough) was reduced by the end of the study, lower in the varenicline compared with the placebo group.

**Table 7A.** Metabolic and Cardiovascular Parameters Measured at Baseline and Week 12.

|  | Baseline<br>mean (SD) |  | Week 12<br>mean (SD) |  | *P* value<br>1 | *P* value<br>2 | *P* value<br>3 |
| --- | --- | --- | --- | --- | --- | --- | --- |
|  | Varenicline | Placebo | Varenicline | Placebo |  |  |  |
| **Weight (Kg)** | 75.5 (13.4) | 78.4 (15.2) | 76.9 (14.2) | 78.8 (15.1) | 0.0742 | 0.9315 | 0.0965 |
| **BMI** | 26.81 (4.52) | 27.76 (4.61) | 27.12 (4.52) | 27.88 (4.03) | 0.1175 | 0.8724 | 0.3434 |
| **SBP (mmHg)** | 126.2 (11.8) | 128.0 (12.7) | 125.1 (9.6) | 126.4 (8.9) | 0.6175 | 0.0864 | 0.8834 |
| **DBP (mmHg)** | 76.1 (8.1) | 77.8 (10.5) | 76.7 (8.6) | 77.20 (9.4) | 0.0904 | 0.7602 | 0.3404 |
| **HR (b/m)** | 74.7 (10.4) | 76.7 (10.8) | 73.8 (8.7) | 75.4 (11.6) | 0.6282 | 0.5288 | 0.3979 |

□ = Baseline vs. Week12 – Varenicline (N=57)
□ = Baseline vs. Week12 – Placebo (N=56)
□ = Varenicline vs. Placebo

**Table 7B.** Metabolic and Cardiovascular Parameters Measured at Baseline and Week 24.

|  | Baseline<br>mean (SD) |  | Week 24<br>mean (SD) |  | *P* value<br>1 | *P* value<br>2 | *P* value<br>3 |
| --- | --- | --- | --- | --- | --- | --- | --- |
|  | Varenicline | Placebo | Varenicline | Placebo |  |  |  |
| **Weight (Kg)** | 74.6 (13.7) | 77.8 (14.2) | 76.8 (14.5) | 78.3 (14.1) | 0.0643 | 0.9415 | 0.0923 |
| **BMI** | 26.56 (4.65) | 27.84 (4.02) | 26.98 (4.56) | 27.93 (4.00) | 0.6468 | 0.9212 | 0.2857 |
| **SBP (mmHg)** | 125.4 (12.1) | 128.2 (12.6) | 124.4 (8.3) | 126.0 (8.2) | 0.6218 | 0.0549 | 0.7022 |
| **DBP (mmHg)** | 76.2 (8.2) | 76.8 (10.1) | 78.0 (8.9) | 77.2 (8.4) | 0.1075 | 0.8451 | 0.3453 |
| **HR (b/m)** | 75.1 (10.4) | 78.2 (11.3) | 74.5 (8.9) | 76.7 (12.2) | 0.7134 | 0.3181 | 0.5787 |

1 = Baseline vs. Week24 – Varenicline (N=51)
2 = Baseline vs. Week24 – Placebo (N=44)
3 = Varenicline vs. Placebo

Measures of urge to vape across the treatment phase of this study were consistently attenuated with varenicline; at week-4, MNWS craving sub-score of 0.56 (0-2) in the varenicline group was significantly lower than 1.61 (0-2.75) (P = .0018) in the placebo group. On the contrary, Varenicline did not attenuate increase in appetite; at week-4, MNWS appetite sub-score of 1.43 (0-3) in the varenicline group was significantly higher than 0.62 (0-1) (P = .0029) in the placebo group.

## DISCUSSION

In our experience and according to recent surveys both youth and adults demonstrate a growing interest in quitting ECs (34-36). In this study, participants’ strong desire to quit vaping was largely due to concern about the potential health risks of long-term EC use and the need to break dependency on vaping products. In spite of the growing interest in vaping cessation, there is no evidence to inform recommendations to assist ECs users intending to stop using their vaping products (37). This RCT is the first to investigate the efficacy and safety of FDA-approved smoking cessation medications for aiding vaping cessation in adult EC users. The findings suggest that varenicline can help EC users to give up vaping and has an acceptable safety profile. Successful vaping cessation with varenicline was also reported in a case report of a 53-year old EC user(38).

At all measure time points, varenicline combined with vaping cessation counseling was superior to placebo. Among EC users, varenicline more than doubled the chance to quit vaping compared with placebo; cotinine level–verified CAR in the varenicline group plus vaping cessation counseling was 40.0% at weeks 4 to 12, and 34.3% at weeks 4 to 24. These results are quite remarkable considering that varenicline only alleviates nicotine withdrawal symptoms and cravings, but cannot replace the need for vaping-related rituals. The ORs for the varenicline group in the present trial were similar than the ORs in previous RCTs of smoking cessation (25,26), NRT cessation (33), and smokeless tobacco cessation (39). These findings suggest that EC users with a relatively high EC dependency (as shown by their previous history of failed quit attempts and relatively high PSECDI score) have similar difficulty quitting compared with smokers in the general population, yet a number do respond to the intervention.

Varenicline is a specific partial agonist and antagonist of the α4β2 nicotinic acetylcholine receptor that has been found to be effective in increasing abstinence rates among cigarette smokers. It is expected to assist EC users quit vaping in light of its mechanism of action that attenuates nicotine withdrawal symptoms and craving (40,41). In line with these observation in cigarette smokers, varenicline was shown to be consistently effective at reducing urge to vape in EC users. Another mechanism by which varenicline facilitates sustained abstinence is by reducing the likelihood of relapse to smoking during a quit attempt (25,26,42). Although relapse prevention was not formally investigated, this effect of varenicline was confirmed in the present study; after stopping varenicline (between week-12 and week-24) vaping relapse rate increased by 17.2% compared to 9.9% after stopping placebo.

Relapse to tobacco cigarette smoking was not observed in this study. Recent work in the US has found that ECs bans have been associated with a substantial increase in cigarette sales and relapse back to smoking (43). However, RCT of vapers with a strong desire to quit are different from research in the context of real-world setting (44).

Presence of cohabitant vapers and high level of anxiety greatly reduced the odds of success for abstinence from vaping, similar to what is observed in cigarette smokers (45). Smokers with anxiety disorders have more severe withdrawal symptoms during smoking cessation than smokers without anxiety disorders and are less likely to quit (46). The presence of smokers in the household is known to be among the strongest sociodemographic predictors of quitting smoking in adult cigarette smokers (47,48). In future, cessation interventions for EC users should take into consideration these specific modifiers.

The safety profile of varenicline in this study was good and similar to that of previous varenicline trials of smokers in the general population (25,26). Nausea, flatulence, and abnormal dreams occurred more frequently in the varenicline group than in the placebo group. As consequence of vaping cessation, the frequency of dry mouth, sore throat, and cough was reduced by the end of the study and much lower in the varenicline compared with the placebo group. No difference in blood pressure and heart rate was noted throughout the intervention. A gradual gain in weight was observed in the varenicline group, but the increase was non-significant compared to placebo.

This RCT has several strengths: 1) use of continuous abstinence rate as a robust primary efficacy endpoint of the study; 2) use of salivary cotinine measurements to objectively verify abstinence from vaping; 3) careful verification of compliance with study medications attained by drug adherence checks; 4) detailed characterization of study participants, that include their vaping patterns, details of their vaping products, and their reasons to quit vaping; 5) use of specific vaping cessation counseling and vaping tapering plan for the study.

Despite these strengths, the study has several limitations. First, findings in a population of adult EC users cannot be extended to young users. Nonetheless, the potential of behavioral interventions for vaping cessation in vapers 18-24 years old has been recently reported (49). Second, findings were restricted to a selected population of participants who had a strong desire to quit ECs, high EC dependency and used almost exclusively refillable vaping products, thus limiting the generalizability of the results. Third, the 6-moth follow-up is limited and longer follow-up should be considered in future studies. Fourth, the impact of vaping cessation counseling could not be assessed as the study was not designed to test the isolated effect of the behavioral intervention.

Future studies with adequate control for the behavioral intervention will be necessary to evaluate the efficacy of this intervention component in EC users. Fifth, a few study participants had their last follow-up visits scheduled during the first wave of the pandemic, which may have affected quit rates. In spite of local lockdown restrictions, participation in the study remained elevated.

This was because the COVID-19 provisions in the Italian ministerial decree allowed people to leave their homes if they could provide proof that they had an appointment at the hospital (subjects in clinical trials were permitted on hospital grounds in full compliance with the anti-COVID rules that restricted hospital access). People used this opportunity to get over the lockdown restriction’s stay-at-home mandate.

Vaping cessation is virtually unexplored and there is a clear need for treatment protocols and guidelines to advance best practice and outcomes for EC users who want to quit. In particular, efficacy and safety of medications approved for smoking cessation by the U.S. FDA for aiding vaping cessation have never been investigated. The findings of the present RCT indicate that inclusion of varenicline in a vaping cessation program for EC users intending to quit may result in prolonged abstinence without serious adverse events. This evidence supports the use of varenicline in cessation programs to help EC users stop vaping and may inform future recommendations by health authorities and healthcare providers. Studies with longer follow-up should be conducted to evaluate long-term efficacy.

## Data Availability

All data produced in this work are contained in the manuscript

## Author Contributions

Pasquale Caponnetto, Paolo Marco Riela, and Carmelo Fabio Longo had full access to all of the data in the study and take responsibility for the integrity of the data and the accuracy of the data analysis.

*Concept and design: Pasquale* Caponnetto, Davide Campagna, Riccardo Polosa.

*Acquisition, analysis, or interpretation of data:* Pasquale Caponnetto, Paolo Marco Riela, Carmelo Fabio Longo, Marilena Maglia, Davide Campagna,

*Drafting of the manuscript:* Pasquale Caponnetto, Davide Campagna, Paolo Marco Riela, Carmelo Fabio Longo, Barbara Busa’, Riccardo Polosa.

*Critical revision of the manuscript for important intellectual content:* Pasquale Caponnetto, Jasjit S. Ahluwalia, Chistopher Russell, Riccardo Polosa.

### Statistical analysis

Paolo Marco Riela, and Carmelo Fabio Longo

### Obtained funding

Pasquale Caponnetto, Riccardo Polosa.

*Administrative, technical, or material support:* Pasquale Caponnetto, Marilena Maglia, Barbara Busa’, Riccardo Polosa.

### Supervision

Pasquale Caponnetto, Davide Campagna, Riccardo Polosa.

## Acknowledgments

We are grateful for the contributions of the Centro per la Prevenzione e Cura del Tabagismo (CPCT, University of Catania, Italy) staff, local vape shops owners, Center of Excellence for the Acceleration of HArm Reduction (CoEHAR, University of Catania, Italy) CoEHAR communication team at the University of Catania, and unpaid interns of the Psychology Section of Dipartimento di Scienze della Formazione (DISFOR) at the same University, who assisted with subjects recruitment.

## Appendix 1

### Vaping cessation counselling

Subjects in both treatment groups received the same vaping cessation counselling throughout the whole duration of the study. One-on-one counselling were provided at each visit for a total of 1015 minutes. Two experienced clinical psychologists delivered this counselling. Our approach to vaping cessation was partially adapted from the 5 ‘s brief tobacco interventions for smokers who are ready to quit (**i**). This behavioral intervention is described in detail below.

<u>First</u>, we collected information about participants’ frequency and intensity of use of vaping products (at baseline) by **asking**:

“Would mind if take a look at your vape/e-cigarette?” “How often do you vape in a day?”

“How much do you normally vape?”

*Depending on the device, questions were modified as:*

*“On average, how many cartridge/pods did you consume yesterday/last week/last month?” “On average, how much e-liquid did you consume yesterday/last week/last month (in ml)?”*

*“What level of nicotine do you use in your vaping product (in mg/ml)?”*

“How soon after you wake up do you first use your vape (i.e. time to first vape)?” “When is the last time you vaped?”

“Why have you decided to quit vaping?” or “What made you decide to quit vaping?”

<u>Second</u>, we **assessed** readiness to quit vaping by asking two questions: ‘‘Do you plan to quit vaping within the next 30 days?’’, ‘‘Do you wish to participate in a vaping cessation program?’’ Before attending the baseline visit (and committing to a target quit date - TQD), potential study subjects were asked to reduce the daily use/consumption of their vaping product by at least 50%.

Depending on the type of vaping product used, we used different definitions to indicate a 50% reduction. If the users was vaping a refilling device, the 50% reduction is indicated by a reduction in e-liquid volume consumed per day (e.g. reducing down to at least 2 ml/day if he/she vaped on average 4 ml/day). If the users was vaping a closed system (e.g. prefilled pod/cartridge) the 50% reduction is indicated by doubling the no. days over which the same volume in consumed (e.g. reducing down to at least 1 pod every two days if he/she consumed 1pod/day). Potential study subjects were instructed to gradually taper down daily consumption at their own pace, over time. When vaping frequency was reduced by 50% (indicating readiness to commit to vaping cessation plan and TQD), they were admitted to the baseline visit.

<u>Third</u>, those who successfully reduced by 50% their daily use were **assisted** with a quit plan (combining vaping reduction, cessation counseling, use of varenicline, and close follow ups). Participants were instructed to set a TQD, ideally within two weeks. This is an important step, but one on which participants must take the lead and choose for themselves because setting a TQD creates accountability and indicates serious commitment. Participants were instructed to increase varenicline dosing prior to the chosen QD according to manufacturer’s recommendations.

Varenicline treatment lasted for 12 weeks. Participants were reminded of the challenges posed by craving and nicotine withdrawal symptoms when stopping vaping products completely and counseled on how to cope with them to avoid a relapse to vaping (or worse to smoking). Close follow up in the first four weeks of the cessation program was arranged to assess participants’ progress, review stress coping skills in order to mitigate the possibility of vaping relapse, address varenicline’s adverse events, and maintain participants’ motivation to quit.

Participants were motivated to stop vaping and therefore in a stage of *determination* to change. s a consequence, our counseling activity aimed to facilitate participants’ transition to the stage of *action* and to keep the desired change when reached (**ii**). This is normally achieved by assisting in dealing with cravings and withdrawal symptoms, and by promptly addressing slips and relapses.

### Assisting in dealing with cravings and withdrawal symptoms

Although there is no formal guidance on how to quit vaping, there is a wealth of information on how to cope with the nicotine withdrawal syndrome that reliably follows cessation of cigarette smoking. As nicotine is an important determinant of e-cigarette dependence, the withdrawal effects experienced upon cessation of vaping may be similar in nature, frequency, and intensity to those experienced when trying to quit tobacco cigarettes. One of the biggest challenges for those who try to quit smoking or vaping is coping with cravings and other nicotine withdrawal effects (**iii, iv, v**). Therefore, vaping cessation counselling may usefully adapt strategies that have long been trained as part of behavioral counselling for smoking cessation.

Practical counseling in this study focused on two elements:

Participants were adviced to use the following coping skills and strategies: a) adopt physical activities, possibly an exercise/sport that is fun for you like playing tennis, ultimate frisbee, taking a swing at batting cages, but even a short walk or dancing can do the trick (the endorphin boost you can get from physical activity can crush a craving); b) use a distraction to take your mind off the craving, by reading a blog/book, playing an instrument, watching TV or a movie, playing a game, doing sudokus or solving crossword puzzles, playing with a pet, texting friends (cravings will pass, if you can give them a minute or two, hence find the activities that will keep you busy for those few minutes); c) throw out anything that reminds you of vaping and that could trigger a craving by creating a temptation-free home/office (make sure your room, backpacks, purses and pockets are free of any e-cigarettes and things you need to vape); d) break behavioral habits, by replacing behavioral habits connected to vaping (e.g. starting the morning by drinking coffee and vaping an e-cigarette) with new rituals (e.g. replace morning coffee with juice or tea). Effective coping strategies may also include controlled breathing exercises (deep/slow breathing), or alternative distraction techniques (e.g. taking a shower, chewing sugar-free gum, eating fresh fruits, fiddling with a pen or squeezing a stress ball, and drinking from a water bottle).

### Promptly addressing slips and relapses

There is very limited information about slips and early relapse when quitting vaping. However, similar to smoking cessation, vaping cessation is not a single event, but rather a process, in which relapse to vaping is a common event. Our approach was partially adapted from recommendations used for assisting smokers who relapse early in the course of their smoking cessation program (**vi**).

Slips and early relapse must be addressed promptly. Participants will have a better sense of their vaping triggers, and counselors can better focus on how to get quickly him/her back on track: “No matter what, you’ve proven that you can do this. Relapses is common, but must be used as a learning experiences through which one can better focus coping skills and improve how to get through the change process”. Slips and early relapses are best addressed by telephone counselling (**vii**). Participants are provided with contact details of their counselor for these situations. It is fundamental to identify the vaping trigger/cue so that the cessation strategy can be individualized with focused coping skills. Moreover, we reviewed whether the subject used the cessation medication in an effective manner and determine whether the drug was helpful.

